# Monitoring of emerging nutritional factors impacting health outcomes: KOMPAS prospective family cohort study

**DOI:** 10.1101/2024.03.03.24303671

**Authors:** Eliška Selinger, Marina Heniková, Martin Světnička, Anna Ouřadová, Monika Cahová, Jana Potočková, Pavel Dlouhý, Dana Hrnčířová, Eva el-Lababidi, Jan Gojda

## Abstract

**Introduction:** The dietary habits of the population are undergone dramatic changes in recent years, with an increasing proportion of people adopting different variants of plant-based diets, limiting their consumption of food of animal origin. Moreover, the shift toward a plant-based diet is supported by scientific reports promoting them as a more sustainable dietary option, which is necessary to adopt on a global scale to mitigate the human influence on climate change. However, despite their growing popularity, many questions regarding their safety and long-term health effects remain unanswered. One of the biggest concerns is the health impact during childhood and adolescence, due to the higher risk of development of severe nutritional deficiencies accompanied by the lack of good quality evidence to guide clinical recommendations and management in those groups. To close the knowledge gap, we decided to establish a family cohort study with increased representation of vegetarian and vegan families with the aim to describe not only adult and child-specific outcomes associated with plant-based diet, but also shared family risks. The aim of the presented manuscript is to introduce the background of the establishment of the family cohort study and provide a description of its protocol.

**Methods:** To investigate the long-term effect of these newly emerging dietary habits, a cohort of families with at least one child under 7 years of age and with the same diet followed by all members was established. The goal for the first year of the study was to recruit at least 40 vegetarian, 40 vegan, and 60-80 omnivore families. At the time of enrollment, biological samples, as well as medical history, together with 3 day dietary records, were taken. The initial active follow-up is planned for at least 5 years.

**Ethics and dissemination:** The study was approved by the Institutional Review Board no. EK-VP/391012. The results of the study will serve as a basis for future research, as well as clinical guidelines and dietary recommendations in Czechia and neighboring regions.

## Introduction

According to the consumer survey in Czechia in 2022, the number of consumers who limit their consumption of animal products was reported to be 39%, an increase of 10% since the last survey conducted in 2020. In more detail, in the population of 18-35 years of age, 3% of consumers are reported to be vegan, 7% vegetarian, 4% pescetarians and 25% flexitarian, which is sharp increase compared to 1% of vegans and 3% of vegetarians in the overall population. ^1^ The increased proportions in younger generations, who are at an age in which they plan to conceive or are already raising children, are a source of a growing medical concern especially due to the lack of epidemiological studies describing the long term health outcomes of plant-based diets introduced during early childhood. Therefore, despite the lack of more precise statistics, it can be assumed that the proportion of children with a decreased amount of animal food in their diet would also be on the rise in the coming years.

However, decreased consumption of animal-based food was associated with a potential increased risk of several nutritional deficiencies. These potential issues could become even more important to consider during sensitive periods of growth like pregnancy or childhood.^2^ With the growing number of families opting for raising their child on a purely plant-based diet or with radically decreased consumption of animal-based food, the questions about the impact of this decision on child growth and health are getting to be more pressing than before. However, there is still a scarcity of research on vegetarian and vegan diet in children population, with only cross sectional studies without the possibility to investigate the medium to long term effect of meat-reducing diet on the growth and development of the children. ^3–5^ This uncertainty is also illustrated by the discrepancy in available national recommendations on plant-based diet, with some countries framing vegan diet as a generally safe option, while others requiring more strict approach, including active discouragement of parents.^6^

To investigate the impact of plant-based diet on health, in adults as well as children, it was decided to establish a pilot family cohort of Czech families following vegan, vegetarian or omnivore diet. The main aim of the study is to establish a basis for future research on a plant-based diet in childhood in the context of central Europe. Part of that effort is also the project is also a creation of a biobank available for future analysis. Moreover, the study should provide medical professionals and nutrition and public health specialists with detailed data on the early development of vegan children. The overall objectives are to recruit 40 vegetarian, 40 vegan, and 60-80 omnivore families, with family defined as two adults living together with at least one child under 7 years of age. The recruitment should take place during the first two years of the project.

## Research priorities

Based on the currently available evidence, which was summarized in previously published umbrella and systematic reviews ^7,8^ accompanied by hand-search of later published studies and based on previous research projects ongoing in Czechia, we have decided to select 3 key research topics, which will be preferably analyzed and investigated in greater detail. Two of those topics are characterized by assumed higher risk of deficiencies in vegetarians or vegans accompanied by higher impact of their nutritional inadequacy during childhood. In contrast to those, one topic was selected as the assumed positive effect of vegetarian or vegan diet, at least in adulthood. Therefore the goal is to investigate if similar potentially positive patterns can be also found and confirmed in childhood. Brief justification for their selection can be found below.

### Bone health

Recently, vegetarian and vegan diets were associated with lower mineral density of bones and increased risk of fractures. ^9–11^ However, the evidence explaining the difference in risk remained inconclusive. As an example, while in the study published in 2007, the increased risk of fractures in vegan group compared to omnivores could be explained by lower calcium intake, with participants consuming at least 525mg/calcium daily having no obvious differences in risk between dietary groups^9^, re-analysis of the EPIC data from 2020 identified higher risk of total as well as site-specific fractures among vegans, even after adjustment for BMI and intake of calcium and protein^10(p202)^. Several risk factors were speculated as possible causes: lower intake of protein, calcium, vitamin D as well as their combination with other nutritional deficiencies or non-dietary lifestyle choices (e.g. differences in physical activity). In line with findings among adults, lower total and lumbar spine BMD z-scores was observed among prepubertal vegetarian children compared to omnivores in polish cross-sectional study involving 53 vegetarian and 53 omnivorous healthy children^12^, with further data on children missing. As any risk to the development and growth of bones during childhood should be taken seriously, investigation of markers of bone health and associated factors is taken as a priority.

### Thyroid health

Despite protein, vitamin B12, vitamin D, iron or calcium being the usual concerns in plant-based diet, recent studies highlighted the potentially overlooked but important issue: iodine.^13–16^ Urinary iodine levels in children were lower in young vegans compared to omnivores^5,17^, even when calculated as proportions to urinary creatine levels^18^. Similar trends were also observed in adults. ^19^ Moreover, vegan and vegetarian diets were also associated with lower intake of selenium or vitamin A, with both having potential negative influence on thyroideal health. Ironically, vegan dietary practices and especially associated consumption of supplements containing kelp was also connected to cases of thyroid dysfunction due to the iodine overdose^20^ and some proportion of over-supplemented children was also recorded in previously published Czech research.^17^ The issue could be also potentially worsened by the limited regulation of dietary supplement market, especially taking into account the significant differences in iodine content in different products freely available without medical consultations. ^21^ Iodine deficiency is a long term issue in Czechia, as the iodine soil content is naturally low. The deficiency in the general population was solved using fortification of salt as well as fortified feed for dairy cows ensuring increased amount of iodine in milk and dairy products, thanks to which the overall population iodine status was brought to healthy normal.^22^ Despite the general success of this public health intervention, some subgroups of population are still at marked risk for iodine deficiency without further support, e.g. pregnant women and newborns.^23^ With the increasing popularity of plant-based diets, families following vegetarian and vegan dietary patterns could become a new high-risk group unreached by current prevention efforts. Research on thyroid status, risk factors and potential means of prevention is therefore warranted.

### Cardiometabolic health

Vegetarian and vegan diets have shown a correlation with favorable cardiometabolic profiles, including reduced cholesterol and triglyceride levels, as well as a potential decrease in the risk of diabetes and cardiovascular events.^7,24–26^ What’s intriguing is the idea that these beneficial effects of a plant-based diet may manifest early in life, even in children. When we consider research suggesting that the early stages of atherosclerosis can begin in childhood, adopting a vegetarian or vegan dietary pattern may offer substantial advantages for cardiovascular health.^27^ However, the available data on lipid profiles in vegetarian and vegan children are inconclusive, with only a limited amount of small and cross-sectional study available.^8^ As non-communicable diseases become the most common causes of mortality and morbidity in high-income countries, with significant burden associated with them in central European region, potential positive impact of shift towards plant-based nutrition was selected as the third research priority.

## Methods

### Design

KOMPAS is a prospective cohort single center study.

### Participants and setting

Participants are invited through advertisement on social media as well as through clinical practice of cooperating pediatricians who are actively inviting families which fulfill the inclusion criteria. Inclusion criteria are defined as being a family consisting of two adults and at least one child under 7 years of age. All family members should follow the same dietary pattern (self-identified vegan, vegetarian or omnivore) and be willing to undergo the examination procedures, including sampling of biological samples. For the purpose of our study, vegan diet is defined as a diet excluding meat and meat products, fish, milk and dairy and eggs. Vegetarian diet is defined by exclusion of meat and meat products together with fish, but consuming eggs or dairy. Omnivore diet is defined as a diet with no strict dietary restrictions put on the food of animal origin. Diet group membership was determined by self-identification. The Exclusion criteria are: different diet followed by different family members, only child older than 7 years, inability or unwillingness to undergo full clinical examination and sampling. From a medical perspective, having disease associated with malabsorption (pancreatitis, PKU etc) was also considered as a reason for exclusion. No other restrictions apply.

### Data collection

At the baseline visit, detailed medical history and socio-demographic surveys are taken, together with biological samples (blood, urine, stool) for all family members.

#### Anthropometric measures

Height and weight is taken for all family members, while muscle strength (maxHGS), blood pressure, body composition (BIA, Nutriguard-M, Data Input GmbH, Tanita MC-780 MA) and waist and hip circumference measured for adults.

All participants would collect the *3-day dietary record* (3DR). 3DR comprises of three non-consecutive days distributed over the two weeks after the first study center visit (2 weekdays and 1 weekend day). Participants are instructed by a trained dietitian on how to keep diet records and receive written instructions, paper diet record forms, and a digital scale. All consumed foods, beverages, and supplements are recorded, if needed (in novel products for instance) they are asked to take photos of their meals and food packaging. Nutrient intakes will be estimated *post hoc* from raw food data (i.e., diet records) using diet assessment software and food composition databases.

#### Biospecimen collection

While blood is sampled at the study center, stool and 24 hour urine collection is collected at home. All participants are instructed on the correct collection procedure for 24-hour urine and stool collection and equipped with the necessary toolkits to ensure high quality of collection. Venous blood sampling is performed on site after an overnight fast (12 hours) from the antecubital vein. After sampling, the specimens are centrifuged and aliquots coded, kept refrigerated, and stored at −80°C during the same morning until being analyzed. Spot urine samples are collected on site using provided containers. All the parameters will be analyzed in a central ISO-certified institutional laboratory.

During the second year of follow-up, repetition of the in person visit at the clinic is planned, with repeated anthropometric measurement and biological sampling. Passive and distance follow-up (via phone or email) will be sustained for additional 3 years at minimum. Graphical representation of the planned course of the study can be found in the **Table 1**.

**Table 1:**
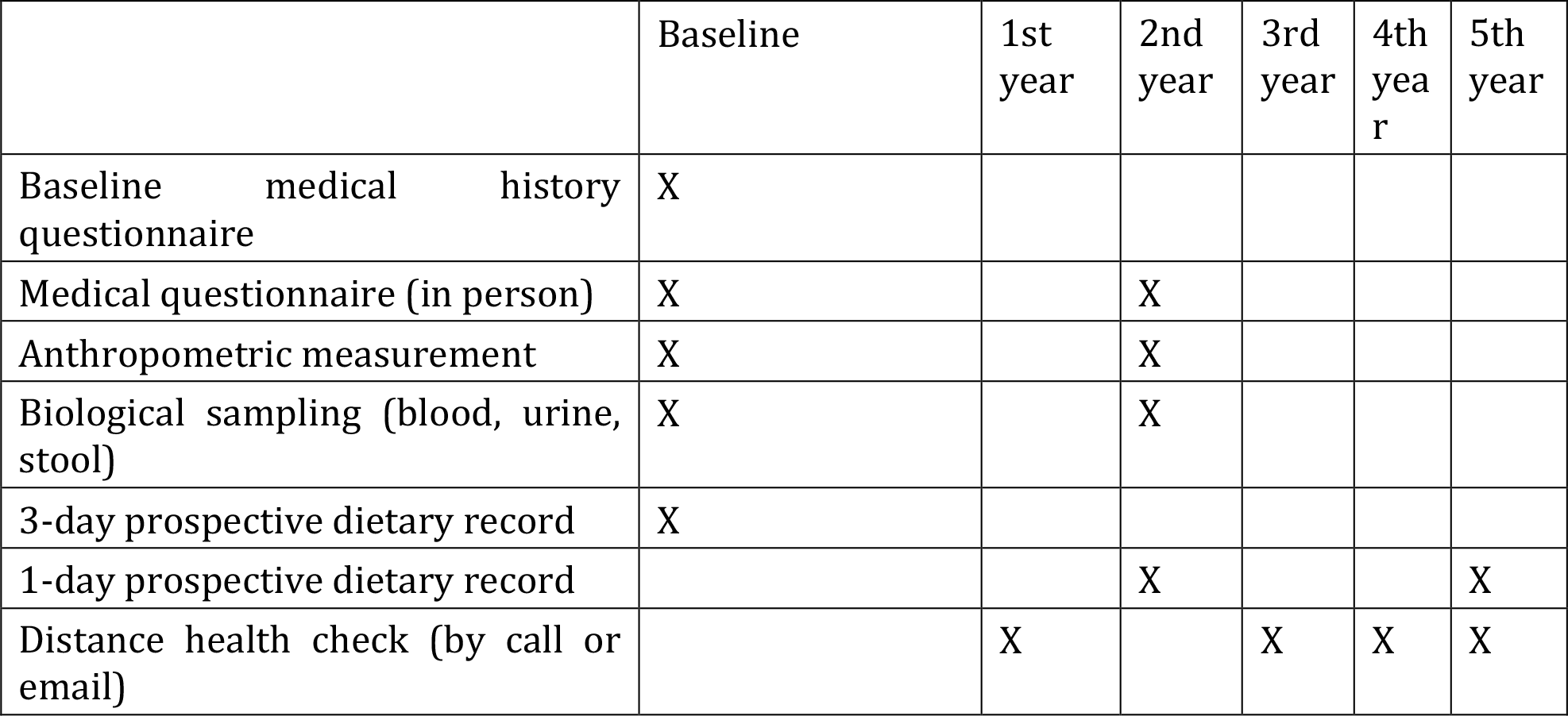
Visualisation of plan of data collection for the first 5 years of follow-up.

|  | Baseline | 1st year | 2nd year | 3rd year | 4th year | 5th year |
| --- | --- | --- | --- | --- | --- | --- |
| Baseline medical history questionnaire | X |  |  |  |  |  |
| Medical questionnaire (in person) | X |  | X |  |  |  |
| Anthropometric measurement | X |  | X |  |  |  |
| Biological sampling (blood, urine, stool) | X |  | X |  |  |  |
| 3-day prospective dietary record | X |  |  |  |  |  |
| 1-day prospective dietary record |  |  | X |  |  | X |
| Distance health check (by call or email) |  | X |  | X | X | X |

From the collected biological samples, a range of markers will be estimated in an accredited institutional clinical laboratory as summarized in **Table 2**. Additionally, the remaining samples will be preserved in a newly established biobank for the use in the future analysis, including metabolomics studies. Stool will be analyzed. Questionnaires used for data collection are available in the **Supplement**.

**Table 2:**
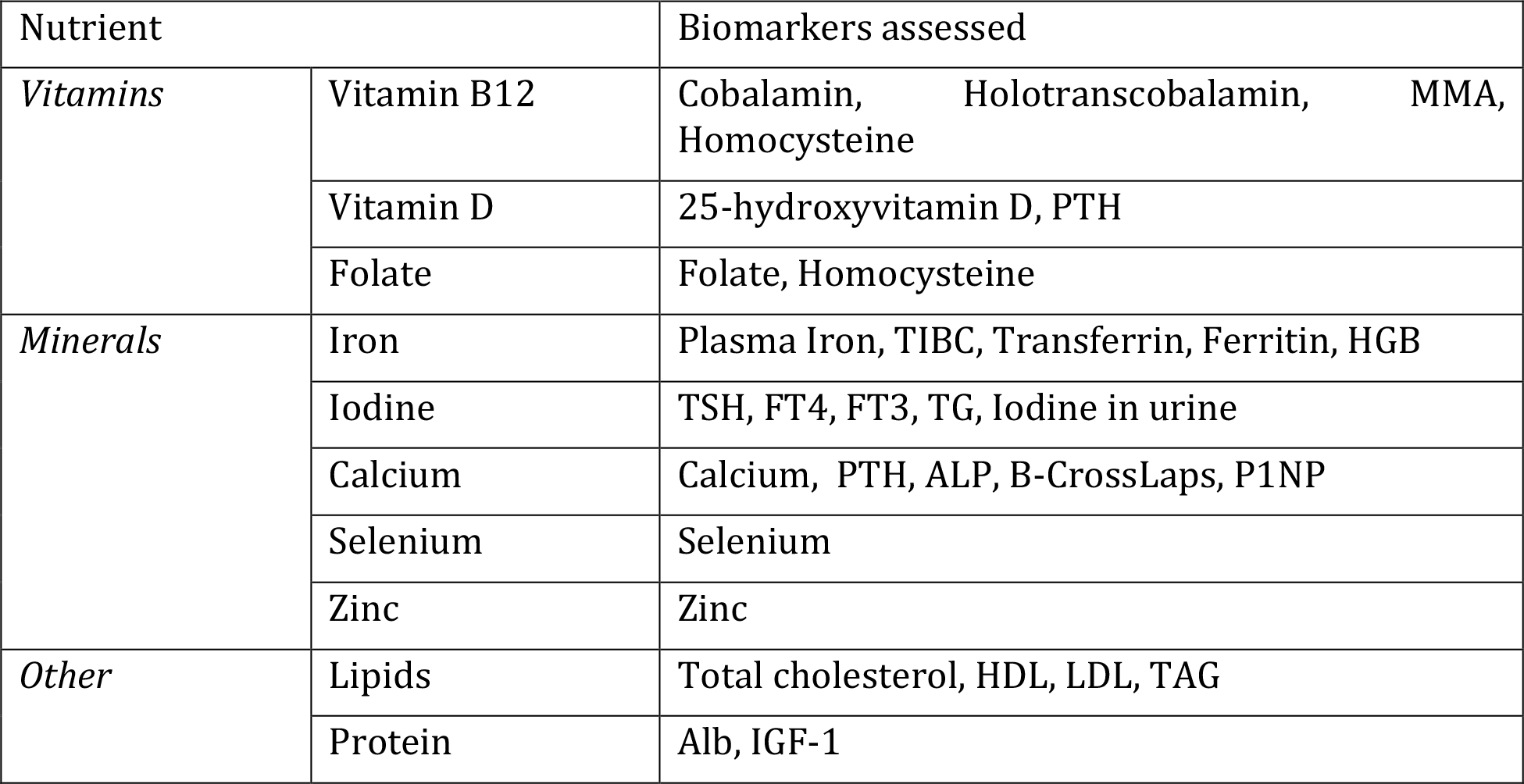
Overview of the main assessed biomarkers related to the risk of prioritized nutritional deficiencies.

### Research questions addressed

Due to the characteristics of the study design, most analyses will be conducted on an exploratory basis. However, several specific research questions will be addressed in studies, utilizing the data collected during the baseline and subsequent follow-up visits. Specific examples of usage of data obtained during the planned visits are described below.

#### Baseline visit

The focus of the analysis of data collected during the first baseline visit will be cross-sectional studies aiming at the description of baseline differences in characteristics of vegan, vegetarian, and omnivore participants and families. From a nutritional perspective, nutrient intake as well as corresponding biochemical markers of nutrient intake will be examined, with special attention given to the nutrients and markers related to pre-selected focus areas (bone health, thyroid health, and cardiometabolic health). See **Table 2**.

#### Follow-up visits

During the second visit, adherence to the baseline diet will be checked and additional biological samples will be taken, opening the possibility to analyze not only changes in dietary intake but foremost the changes in relevant biomarkers and anthropometric measures. The second is especially important for the children cohort, as questions about the healthiness of the growth of vegetarian and vegan children are still raised in many clinical guidelines and professional statements^28^ Due to the rapid rate of growth of children sensitive to the insufficient intake of energy or nutrients in the targeted period, any significant growth issues caused by the inadequate diet could be apparent even after the short term period of follow-up.

### Sample size

The sample size was established mostly as a convenient sample based on previous research, prevalence of vegan and vegetarian eating practices in Czechia and usual practice in the field of vegetarian and vegan children research. Although modest in scale, it holds the potential to reveal essential distinctions and facilitate hypothesis generation for future research. Two main limitations had to be taken into account during sample size planning. First, it is essential to bear in mind the limited prevalence of vegan families within the Czech Republic and the associated challenges in recruiting participants. While veganism being on a rise, the prevalence of fully vegan families is still substantially lower. Moreover, our approach takes into account the reluctance of omnivore families to subject their small children to blood sampling confirmed by previous research organized in the study center. Whole vegetarian and vegan families could be motivated to participate by their perceived higher risk of nutritional deficiencies as well as general unavailability of specialized primary care suited to their needs, for omnivore families neither of those motivators exists. Despite these limitations, the cohort can provide valuable insights into the dietary practices of vegetarian and vegan families within our unique research environment.

### Data analysis

Baseline characteristics of all participants will be described. Between-group differences will be reported as mean differences and 95% confidence intervals, or differences in proportion and 95% confidence intervals, with potential calculation of effect size, where appropriate. The level for statistical significance will be set up at 0,05. When possible, linear or logistic regression adjusting for the main potential confounding variables will be used as part of the statistical analysis. To take into account within-individual as well as between-individual variation, linear mixed models will be conducted in investigation of follow-up results. Data analysis will be conducted by trained epidemiologists using RStudio software [15]. All publications presenting the results will respect the current reporting guidelines.

### Bias considerations

Two types of bias typically appear in the course of nutritional studies and need to be taken into account in their planning: selection bias and measurement bias. It is known that nutritional studies, even observational, have low levels of compliance, due to the time demanded for visits and dietary records. In the case of the presented project, the situation is worsened by the involvement of whole families, including young children, and the need to collect not only questionnaire data but also biological samples. The second is especially challenging in children as for them the sampling is significantly more uncomfortable or even distressing. Despite the blood sampling being considered generally safe and having theoretical potential to uncover unknown medical issues, many parents are understandably hesitant to approve such a procedure in children when not necessary from a medical perspective. Resulting situation is that only highly motivated families, which are unusually interested in dietary monitoring or have known medical issues, can be willing to participate. Ironically, in the context of vegan and vegetarian studies, this selection bias can be considered an advantage. It is known that vegan and vegetarians have higher interest in a nutritional and healthy lifestyle^29^, together with high motivation to participate. This interest can create confounding in the analysis of any differences between dietary groups. Selection of highly motivated omnivores therefore can decrease the differences found in comparison to the general population.

Measurement bias is based mainly on the impossibility to obtain objective records of dietary intake. As most of the information recorded is based on self-reporting, it needs to be considered that many participants’ records will not be precise or truthful for a wide range of reasons. To diminish this bias, all medical data are collected using unified questionnaire and trained medical staff. Most importantly, all participants will be educated by a trained dietitian on how to correctly record their dietary intake, including the possibility to add photos of the portions or food products consumed. In case of any doubts in the records, participants are contacted by the dietitian to resolve the issues. Moreover, as biological samples are taken together with the dietary record, it is possible to base any analysis on objective measurements of levels of markers of nutrient intake, together with self-recorded information.

### Ethical considerations

Ensuring the safety and well-being of the participants, including participating children, was taken as a top priority during the planning process and recruitment. Careful steps to create a secure and supportive environment during the study were undertaken, making sure the children are as comfortable as possible during the whole course of the research. The study is conducted according to the guidelines of the Declaration of Helsinki, and approved by the Ethics Committee of Faculty Hospital Královské Vinohrady (protocol code EK-VP/58/0/2019).

### Patient and public involvement

Research team has established collaborative partnerships with vegan and vegetarian organizations, fostering a strong connection with the National Institute of Public Health and national healthcare professional organizations. research team members regularly engage in public conferences, stakeholders meetings and other relevant events where they communicate ongoing study results with a wider audience. Furthermore, publication of nutritional recommendations focused on the needs of vegetarian and vegan citizens is planned, with the goal of ensuring that the knowledge and insights gained from the study benefit not only the research community but also the general public.

## Discussion and conclusion

According to available estimates, over half of the habitable land is used for the purpose of agriculture, with over 70% of that being consumed for the purpose of animal industry, mostly due to the production of animal feed. Moreover, production of animal based food is associated with the production of a significant higher amount of greenhouse gas emissions compared to plant-based products and is in general associated with higher demand for water, as well as more negative impact on eutrophication or deforestation.^30,31^ Due to this, changes in current diets consisting of a significant decrease in consumed amount of animal based food were suggested as a potential way to diminish the negative impact of climate health.^32,33^

Besides environmental reasons, many people are opting for a plant-based diet as a means to prevent the development of diet-related non-communicable diseases, or even as a part of therapeutic regimes. It is assumed that a plant-based diet can decrease the risk of diabetes, cardiovascular diseases, or even some types of cancer.^26^ Previous research on adults has also shown that vegan diet, as the most strict variant of plant-based diet, is associated with lower BMI, better cardiometabolic profile and potentially also lower cancer mortality. Evidence of a potential beneficiary effect of plant-based diet was described in previous research even among children.^27,34^

Last but not least, the most common reasons to decrease the consumption of animal-based food are ethical concerns, especially among vegan families. In contrast to vegetarianism, which has a long history in many cultures and is associated with many religious practices, veganism was created as a new philosophical construct in 1940s. The definition, still promoted by Vegan Societies, describe veganism as “*a philosophy and way of living which seeks to exclude—as far as is possible and practicable—all forms of exploitation of, and cruelty to, animals for food, clothing or any other purpose*”^35,36^. The reasons for this philosophical position is an underlying opinion by which humans should not have treat the non-human animals as inferior and should respect that non-human animals are independent entities capable of feeling and perceiving. Moreover, realities of current mass production of animal based food are often cited as reasons for abstaining from animal based products. As such, veganism is a judicially endorsed worldview protected by the European Convention on Human Rights. ^37^ From a purely dietary perspective veganism means the practice of excluding all food of animal based origin, mainly meat and meat products, including gelatine or bone broth, fish and fish products, milk and dairy and eggs.

Concerns for animal rights are the most common reasons for exclusion of animal based food, confirmed by qualitative as well as quantitative research. Vegetarians and vegans cite animal welfare as most common reasons while they followed selected diet, while taste and enjoyment was more common for omnivores.^38^ Similarly, animal-rights and ecological concerns predicted adherence to vegetarianism, compared even to semi-vegetarianism. ^39,40^ Differences in motivation are also associated with views and tension between different vegetarian subgroup and meat-eating majority, with both vegetarians and vegans in general preferring people adopting the veg(etari)an diet for ethical motivations (animal or environmental).^41^ In contrast to vegetarian groups, non-vegetarians cited health as the main motive to consider adopting vegetarian diet. ^42^ Interestingly, motivation was also correlated with the diet quality among vegetarians, with health motivated vegetarians having higher quality of diet as measured by Healthy eating Index than vegetarians motivated by other reasons.^43^

In contrast to the growing evidence on the importance of the population-wide shift toward plant-based dietary patterns and a necessity to limit the global consumption of animal-based food, especially red and processed meat, reasons for skepticism still prevail. Plant-based diet, especially a vegan diet, has been associated with a wide range of health issues with the most commonly discussed being the risk of development of several nutritional deficiencies, like vitamin B12, vitamin D, vitamin K, vitamin B2, vitamin B6, iron, zinc, selenium, iodine or calcium.^19,44,45^ Recent studies also described an increasing risk for fractures, lower bone density or increased risk of hemorrhagic stroke in vegetarian and vegan populations. ^24,46^ The presented study should therefore bring evidence on the nutritional risks associated with the shift towards plant-based in a widely overlooked population and serve as a basis for modern dietary recommendations on the adoption of vegetarian and vegan diet in the context of central Europe.

## Data Availability

Data are not publicly available.

## Acknowledgements

The work is supported by Ministry of Health grant no. NU21-09-00362.

